# Bulk and single-cell transcriptomics identify tobacco-use disparity in lung gene expression of ACE2, the receptor of 2019-nCov

**DOI:** 10.1101/2020.02.05.20020107

**Authors:** Guoshuai Cai

## Abstract

In current severe global emergency situation of 2019-nCov outbreak, it is imperative to identify vulnerable and susceptible groups for effective protection and care. Recently, studies found that 2019-nCov and SARS-nCov share the same receptor, ACE2. In this study, we analyzed five large-scale bulk transcriptomic datasets of normal lung tissue and two single-cell transcriptomic datasets to investigate the disparities related to race, age, gender and smoking status in *ACE2* gene expression and its distribution among cell types. We didn’t find significant disparities in *ACE2* gene expression between racial groups (Asian vs Caucasian), age groups (>60 vs <60) or gender groups (male vs female). However, we observed significantly higher *ACE2* gene expression in former smoker’s lung compared to non-smoker’s lung. Also, we found higher *ACE2* gene expression in Asian current smokers compared to non-smokers but not in Caucasian current smokers, which may indicate an existence of gene-smoking interaction. In addition, we found that *ACE2* gene is expressed in specific cell types related to smoking history and location. In bronchial epithelium, *ACE2* is actively expressed in goblet cells of current smokers and club cells of non-smokers. In alveoli, *ACE2* is actively expressed in remodelled AT2 cells of former smokers. Together, this study indicates that smokers especially former smokers may be more susceptible to 2019-nCov and have infection paths different with non-smokers. Thus, smoking history may provide valuable information in identifying susceptible population and standardizing treatment regimen.

## Introduction

In the past two decades, pathogenic coronaviruses (CoVs) have caused epidemic infections, including the server acute respiratory syndrome (SARS)-CoV outbreak in 2003, the Middle East Respiratory Syndrome Coronavirus (MERS-CoV) outbreak in 2012 and the current Wuhan 2019 Novel Coronavirus (2019-nCov) outbreak. They typically affect the respiratory tract and cause severe respiratory illnesses. We have learned from SARS-Cov and MERS-Cov that human populations showed disparities in susceptibility to these viruses. For example, epidemiology studies found that males had higher incidence and mortality rates than females.^1,2^ We believe that the susceptibility to the novel 2019-nCov is also different among population groups. In current severe global emergency situation of 2019-nCov outbreak, it is imperative to identify vulnerable and susceptible groups for effective protection and care.

Recently, Xu et.al. computationally modelled protein interactions and identified a putative cell entry receptor of 2019-nCov, angiotensin-converting enzyme 2 (ACE2), which is also a receptor for SARS-nCov.^3^ Zhou et.al. further confirmed this virus receptor in the HELA cell line.^4^ Interestingly, Zhao et al. found *ACE2* is specifically expressed in a subset of type II alveolar cells (AT2), in which genes regulating viral reproduction and transmission are highly expressed.^5^ They also found that an Asian male had much higher ratio of *ACE2*-expressing cell than other seven white and African American donors, which may indicate the higher susceptibility of Asian. However, the sample size was too small to draw conclusion on this racial disparity. Here, we analyzed four large-scale bulk transcriptomic datasets of normal lung tissue to investigate the disparities related to race, age, gender and smoking status in *ACE2* gene expression. Also, we analyzed two lung tissue single-cell transcriptomic datasets to investigate the distribution of *ACE2* gene expression among cell types, which will provide new knowledge for understanding the mechanism and population disparities of 2019-nCov infection.

## Methods

### Bulk transcriptomics

Two RNA-seq datasets and two DNA microarray datasets from lung cancer patients were analyzed in this study, including a Caucasian RNA-seq dataset from TCGA (https://www.cancer.gov/tcga), an Asian RNA-seq dataset from Gene Expression Omnibus (GEO) with the accession number GSE40419^6^, an Asian microarray dataset from GEO with the accession number GSE19804^7^ and a Caucasian microarray dataset from GEO with the accession number GSE10072^8^. Both RNA-seq datasets were generated with the Illumina HiSeq platform and both microarray datasets were generated with the Affymetrix GeneChip Human Genome U133 Array. The details and processing of data were described in our previous study^9^. All these datasets contain samples from tumor and normal pairs and we only use the normal samples in this study. In addition, we analyzed a GSE34450^10^ microarray dataset of gene expression from small airway epithelium and large airway epithelium of 50 healthy nonsmokers and 71 healthy smokers. In total, 54 samples in the TCGA dataset, 77 samples in the GSE40419 dataset, 60 samples in the GSE19804 dataset, 33 samples in the GSE10072 dataset and 121 samples in the GSE34450 dataset were analyzed. We studied the Reads Per Kilobase per Million mapped reads (RPKM) values for RNA-seq data and Robust Multi-Array Average (RMA)^11^ values for microarray data. All data were log2 transformed to improve normality. The means of data values across samples in datasets from the same platform were highly correlated (Pearson correlation coefficient r=0.9 for microarray datasets and r=0.97 for RNA-seq datasets, Fig. S1), indicating no significant system variation in datasets from the same platform.

Simple linear regressions were used to test the association of *ACE2* gene expression with each single variable of age, gender, race and smoking status. And, multiple linear regression was used to test the association of *ACE2* expression with multiple factors (age, gender, race, smoking status and data platform). Also, ordinal regression was performed to investigate the association between *ACE2* expression and ordinal categorical smoking history. All data management, statistical analyses and visualizations were accomplished using R 3.6.1.

### Single-cell transcriptomics

Two single-cell RNA sequencing (scRNA-seq) datasets available in GEO with accession numbers GSE122960^12^ and GSE131391^13^ were downloaded and analyzed. The GSE122960 dataset was generated from lung tissue of 8 lung transplant donors (including 1 Asian former smoker, 2 Caucasian current smoker and 5 African American non-smokers), using 3’ V2 chemistry kit on 10x Genomics Chromium single cell controller. The GSE131391 study profiled bronchial epithelial cells from 6 never and 6 current smokers using CEL-Seq^14^. Counts of single cells were downloaded, and subsequent data analyses were performed using the Seurat 3.0 package^15^, including data normalization, high variable feature selection, data scaling, dimension reduction and cluster identification. We also used SCANNER^16^ to assist the data visualization and cell type identification.

## Results

### No observed disparities between race, age or gender groups

Inconsistent with the study of Zhao et al.^5^, we observed no significant difference in *ACE2* expression in Caucasian lung tissue samples compared to Asian lung tissue samples in the RNAseq datasets (*p*-value=0.45, Fig 1A). In the microarray datasets, a higher *ACE2* expression was observed in Caucasian samples compared to Asian samples (*p*-value=0.001, Fig 1A). Given that the GSE19804 microarray study focused on female non-smokers while the GSE10072 dataset includes samples from both males and females and both smokers and non-smokers, we believe that the observed disparity may be due to other factors other than race, such as smoking, gender and unknown factors. Therefore, we performed multiple linear regression on multiple independent variables (age, gender, race, smoking status and platform) and found no significant difference between racial groups (*p*-value=0.36, Fig. 1B).

**Figure 1.**
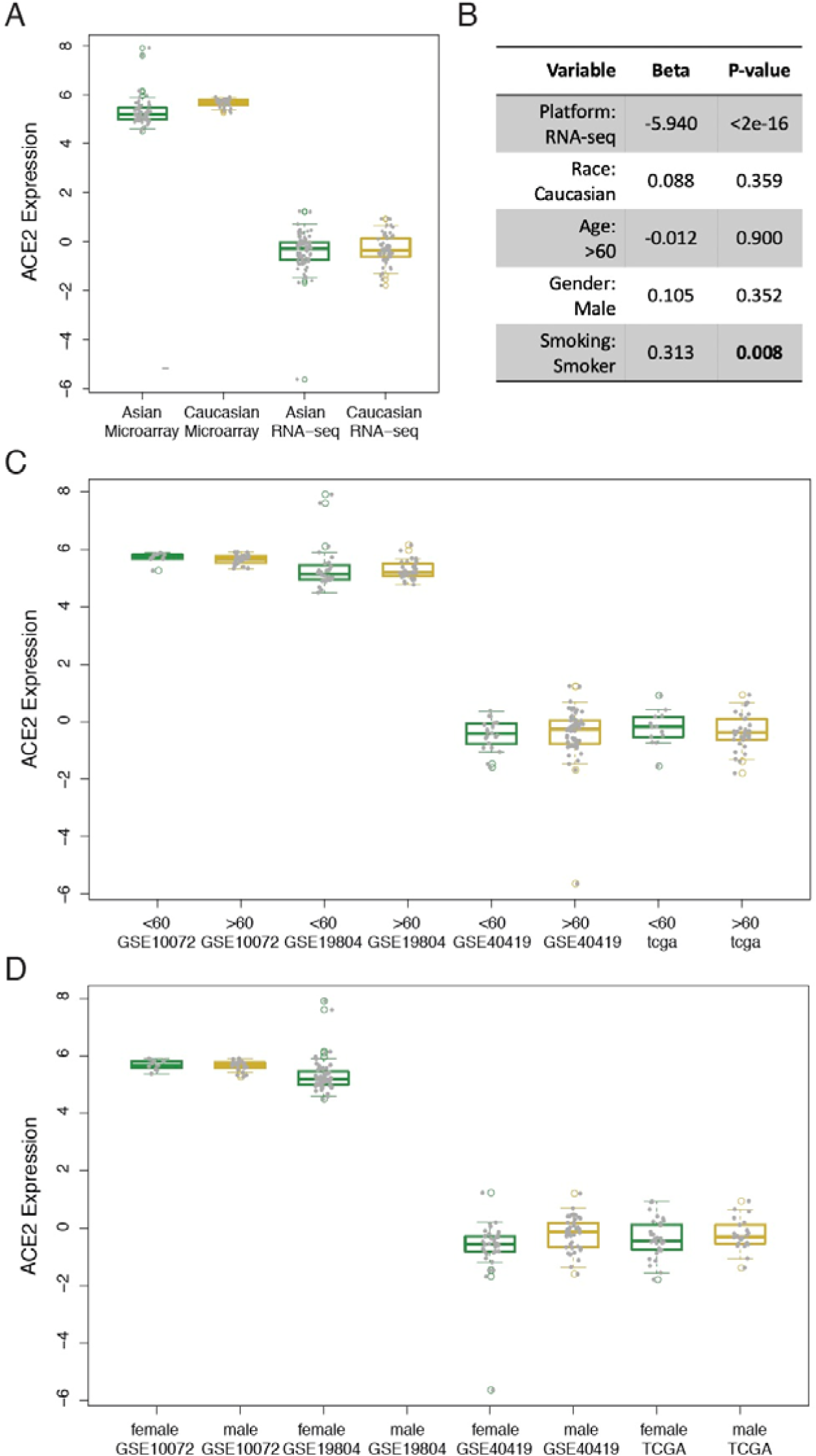
*ACE2* gene expression profiles in groups. **A, C** and **D** shows groups in race (Caucasian vs Asian), age (>60 vs <60) and gender (male vs female). **B** shows the result from multivariate analysis with all factors including age, gender, race, smoking and platforms.

Furthermore, we didn’t observe a disparity between age groups (>60 vs <60) or gender groups (male vs female) in *ACE2* gene expression in each available dataset (Fig. 1C, D). Consistently, multivariate analysis didn’t detect a significant difference between groups of age or gender after other variables (age/gender, race, smoking status and platforms) were adjusted (*p*-value=0.90 for age, *p*-value=0.35 for gender, Fig. 1B). We also consistently found no difference between male and female healthy lung tissue samples from GTEx^17^ (Fig. S2).

### Smokers especially former smokers have upregulated *ACE2* in lung

We found a significant higher *ACE2* gene expression in smoker (including current smoker and former smoker) samples compared to non-smoker samples in the TCGA (*p*-value=0.05) and GSE40419 RNA-seq datasets (*p*-value=0.01, Fig. 2A). Smokers in GSE10072 showed a higher mean of *ACE2* gene expression than non-smokers. The difference is not significant (*p*-value=0.18), which may be due to the small sample size of this study (n=33) with insufficient power to detect the difference. The GSE19804 data which has only non-smoker samples available was not included into the analysis. Adjusted by other factors (age, gender, race and platforms) in multivariate analysis, smoking still shows a significant disparity in *ACE2* gene expression (*p*-value=0.01, Fig. 1B). These data were from the normal lung tissue of patients with lung adenocarcinoma, which may be different with the lung tissue of healthy people. Therefore, we also analyzed a gene expression dataset of airway epithelium from healthy smokers and healthy non-smokers. Consistently, we observed a significant upregulation of ACE2 gene expression in both large airway epithelium and small airway epithelium of smokers (*p*-value=4.99E-4 and 7.02E-3, respectively, Fig. 2C). Furthermore, we investigated ordinal categorized smoking history (non-smoker, former smoker quit more than 15yrs, former smoker quit less than15yrs, and current smoker) and showed results in Figure 2B. In the TCGA dataset, we found a significant trend of *ACE2* gene expression regulation associated with the smoking history that current smokers had the highest expression, non-smokers had the lowest expression and former smokers had that in-between (ordinal regression *p*-value=0.01, Fig. 2B). We found a similar but non-significant trend in the GSE10072 due to its small size (ordinal regression *p*-value=0.11). We didn’t observe such a tread in the TCGA dataset. Instead, we found a higher average expression in recent quitters (<=15 years) compared to non-smokers, current smokers and former smokers who have quit for longer durations (>15 years). This may indicate a difference in *ACE2* gene expression between Caucasian and Asian current smokers, but it is not statistically significantly detected due to the limited sample size in this study (p-value=0.43). Compared to non-smokers, multivariate analysis on all data showed a significant higher *ACE2* expression in former smokers (p-value=0.04) and a higher mean of *ACE2* expression in current smokers, which did not reach statistical significance from current analysis though (p-value=0.11).

**Figure 2.**
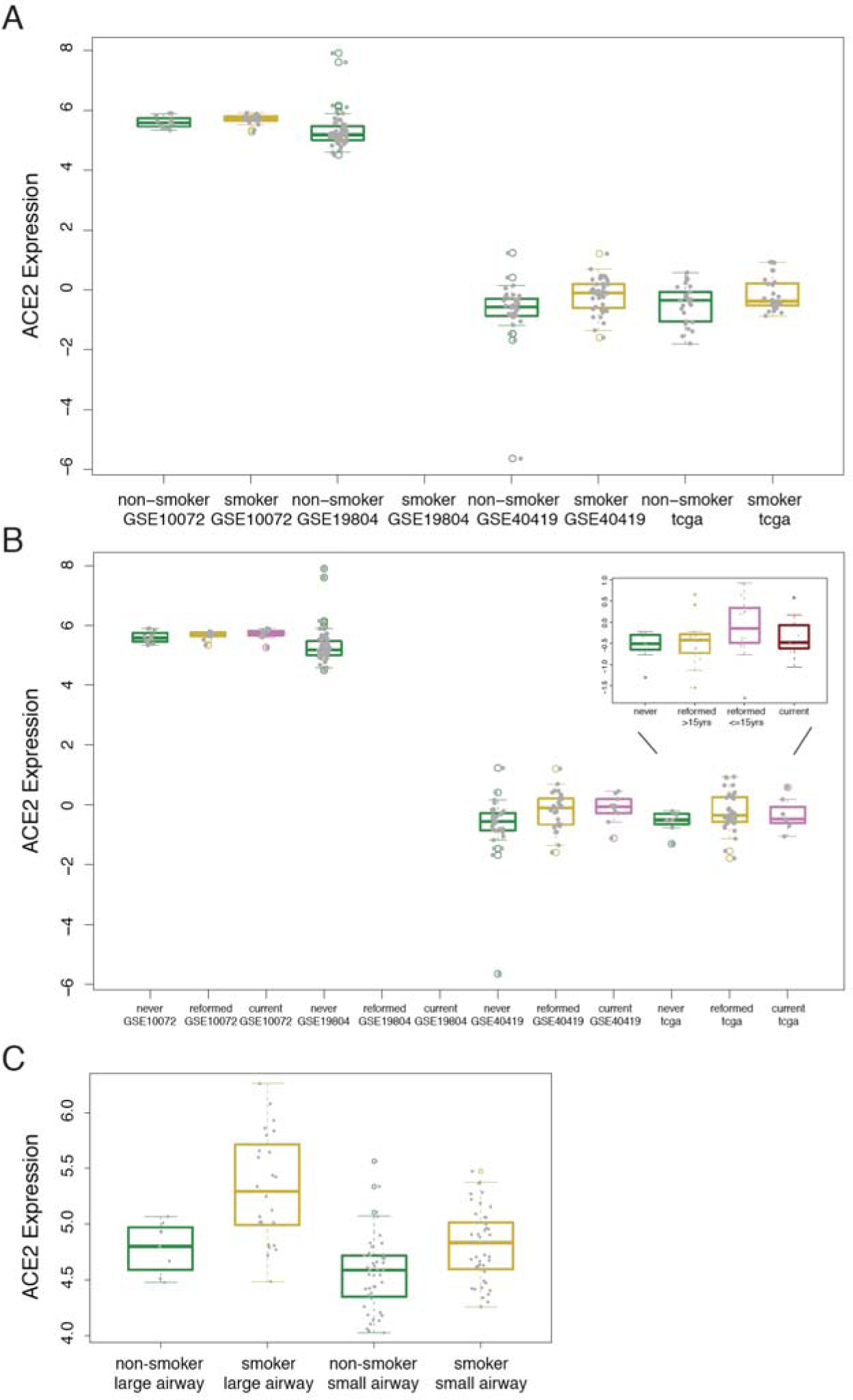
*ACE2* gene expression profiles in smoking groups. **A** shows expression in normal lung tissues of smoker and non-smoker with lung adenocarcinoma. **B** shows expression in normal lung tissues of never-smoker/non-smoker, reformed/former smoker and current smoker with lung adenocarcinoma. TCGA dataset has more categories of smoking history, including never-smoker, smoker reformed more than 15 years, smoker reformed less than 15 years and current smoker. **C** shows expression in healthy lungs of smoker and non-smoker.

### Smokers have upregulated *ACE2* in remodelled cell types

Duclos G et.al. studied human bronchial epithelial cells using single-cell RNA sequencing and found smokers showed a remodeled cell composition in bronchial epithelium with a loss of club cells and extensive hyperplasia of goblet cells.^14^ We confirmed their analysis in this study based on the same set of cell makers, including *KRT5* for basal cells, *FOXJ1* for ciliated cells, *SCGB1A1* for club cells, *MUC5AC* for goblet cells and *CD45* for WBCs (white blood cells) (Fig. 3A,C, Fig. S3). We also identified a smoking related basal cell subpopulation which might be pro-goblet precursor cells and has been discussed in the study of Duclos G et.al.^14^ Interestingly, we found *ACE2* is mainly expressed in goblet cells in current smokers and club cells in non-smokers (Fig. 3B), indicating 2019-nCov may infect different cell types in bronchial epithelium of current smokers and non-smokers.

**Figure 3.**
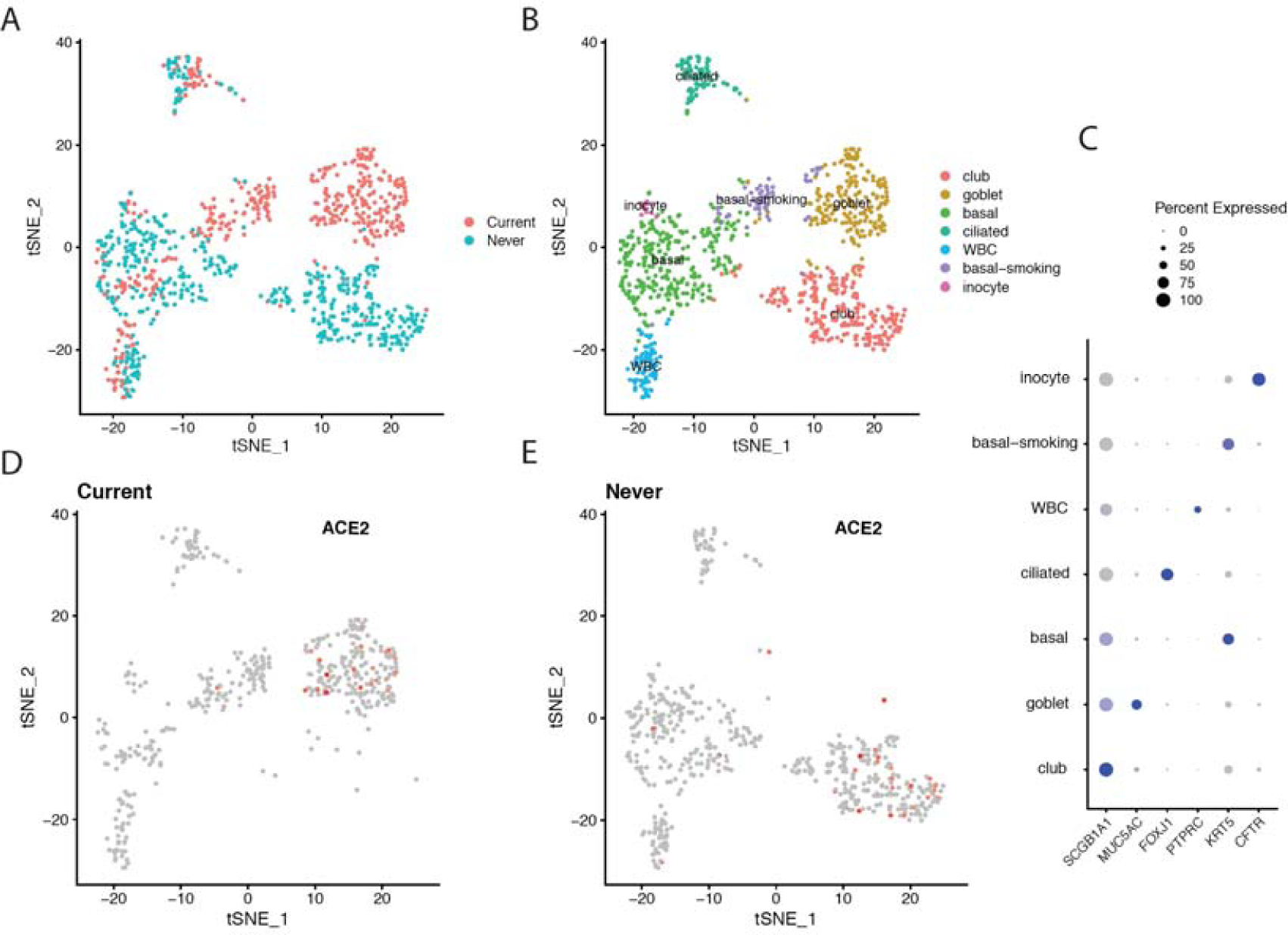
single-cell transcriptomics of bronchial epithelium cells. **A** shows tSNE plots of single-cell transcriptome profiles from never smokers and current smokers. **B** shows identified cell types. **C** shows detection rates of markers in each cell cluster. **D** and **E** show *ACE2* expression in cells of current smokers and never smokers separately.

Further, we analysed a scRNA-seq dataset of whole-lung tissue. Based on the expression of a set of markers shown in Figure 4C and Figure S4, we identified 13 distinct cell populations including alveolar type I (AT1) cells, alveolar type II (AT2) cells, endothelial cells, ciliated cells, club cells, fibroblast, monocytes, macrophages, B, T cell or natural killer T cell (T/NKT), dendritic cells, a former smoker-specific subpopulation of AT2 cells (AT2-reformed) and a current smoker-specific subpopulation of AT2 cells (AT2-smoking) (Fig. 4A). Significantly, current smoker and former smoker showed differently remodelled AT2 cells (AT2-smoking and AT2-reformed, respectively). Also, we found *ACE2* is most actively expressed in AT2-reformed cells in former Asian smokers but not in Caucasian current smokers and African American non-smokers (Fig. 4B, D), which is consistent with our finding from above large-scale bulk transcriptome analysis. In addition, we observed an expression of *MUSC5AC* (the marker of goblet cells) in a subpopulation of club cells of current smokers, indicating the smoking related tissue remodelling with club cell loss and goblet cell hyperplasia (Fig. S5 top). This is consistent with our finding in bronchial epithelium. However, we failed to observe an active expression of *ACE2* in the goblet cells of current smokers in a way similar to what we found in bronchial epithelial cells (Fig. S5 bottom). The limited cell number of this cluster in this dataset might be the reason.

**Figure 4.**
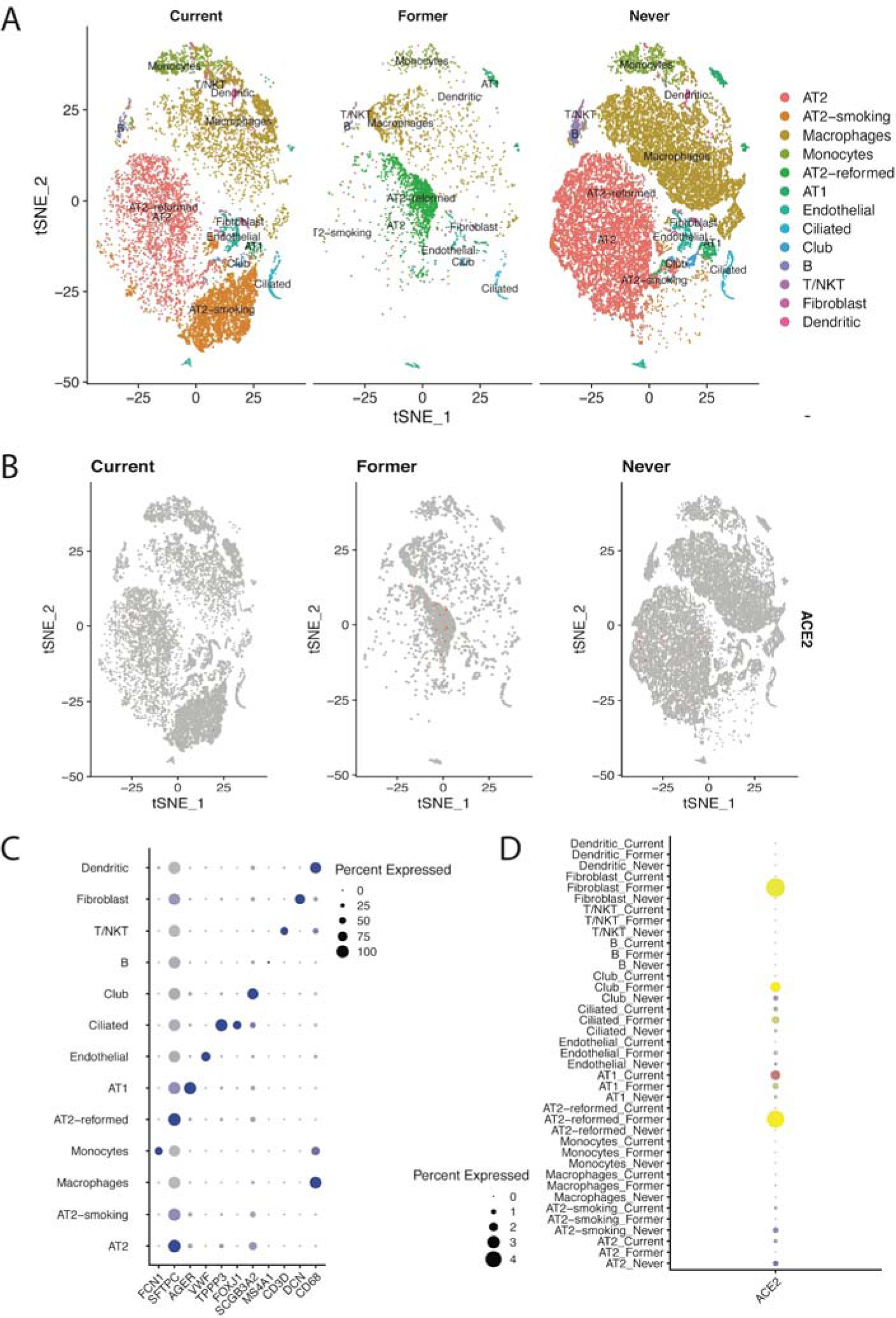
single-cell transcriptomics of lung cells. **A** shows tSNE plots of single-cell transcriptome profiles and identified cell types from never smokers, former smokers and current smokers. **B** shows *ACE2* expression in cells from never smokers, former smokers and current smokers separately. **C** shows detection rates of markers in each cell cluster. **D** shows detection rates of *ACE2* in each type of cells from never smokers, former smokers and current smokers.

## Discussion

In this study, we investigated the disparities related to race, age, gender and smoking status in *ACE2* gene expression by analyzing bulk and single-cell transcriptome data. We found significantly higher *ACE2* gene expression in lung tissue of former smokers compared to that of non-smokers. This may explain the reason why more males (56% of 425 cases) were found in a recent epidemiology report of 2019-nCov early transmission by China CDC^18^. It also consistent with the epidemiology study of 24,554 cases that former smokers (49%) have the higher risk to develop severe disease than non-smokers (14.5%) and current smokers (21.7%)^19^. We didn’t observe significant disparities in *ACE2* gene expression between racial groups (Asian vs Caucasian), age groups (>60 vs <60) or gender groups (male vs female). However, Asian current smokers may have higher *ACE2* gene expression than Caucasian current smokers. The different is not statistically significant in this study but may indicate an existence of gene-smoking interaction.

We also found that *ACE2* gene is expressed in specific cell types related to smoking history and location. In bronchial epithelium, ACE2 is actively expressed in goblet cells of current smokers and club cells of non-smokers. In alveoli, ACE2 is actively expressed in remodelled AT2 cells of former smokers. This may indicate that 2019-nCov infect respiratory tract through different paths in smokers, former smokers and non-smokers, and this may partially lead to different susceptibility, disease severity and treatment outcome.

One limitation of this study is that the small sample size of current single-cell transcriptome datasets has limited power in studying multiple factors involved in this question.

Whether *ACE2* is the only or major receptor of 2019-nCov is unknown. The reason(s) for the tobacco-related disparity in *ACE2* expression is unknown. Studies found smoke significantly increased *ACE2* expression in the lung of rats^20^ and cigarette smoke exposure increased pulmonary ACE2 activities in mice^21^. Controversially, other studies showed chronic cigarette smoke and nicotine decreased *ACE2* expression in rats^22,23^. Thus, substance other than nicotine might contribute to the smoking-related upregulation of *ACE2* found in this study. Further studies are required to find the answer. Despites current limited knowledge, this study indicates that smokers especially former smokers may be more susceptible to 2019-nCov and have infection paths different with non-smokers. Thus, smoking history may provide valuable information in identifying susceptible population and standardizing treatment regimen. Wuhan, stay strong.

### Ethical oversight

There is no direct involvement of human subjects in this study. All the data use existing de-identified biological samples and data from prior studies. Therefore, ethical oversight and patient consent were not handled in this project.

## Data Availability

Two RNA-seq datasets and two DNA microarray datasets from lung cancer patients were analyzed in this study, including a Caucasian RNA-seq dataset from TCGA (https://www.cancer.gov/tcga), an Asian RNA-seq dataset from Gene Expression Omnibus (GEO) with the accession number GSE40419, an Asian microarray dataset from GEO with the accession number GSE19804 and a Caucasian microarray dataset from GEO with the accession number GSE10072. In addition, we analyzed a GSE34450 microarray dataset of gene expression from small airway epithelium and large airway epithelium of 50 healthy nonsmokers and 71 healthy smokers. Also, two single-cell RNA sequencing (scRNA-seq) datasets available in GEO with accession numbers GSE12296011 and GSE13139112 were downloaded and analyzed.

## Supplementary File

**Figure S1. Correlation of four datasets**.

Lower panel shows pairwise scatter plots of data mean across samples in each dataset. Upper panel shows their corresponding Pearson correlation coefficients.

**Figure S2. *ACE2* gene expression in GTEx female and male lung tissues**.

y-axix shows the log10 scaled RNA-seq Transcript Per Million (TPM) values.

**Figure S3. Expression profiles of bronchial epithelium cell markers.**

**Figure S4. Expression profiles of lung tissue cell markers**.

**Figure S5. Expression profiles of *MUSC5AC* (top) and *ACE2* (bottom)**.

